# Plasma biomarkers of Alzheimer’s disease predict cognitive decline and could improve clinical trials in the cognitively unimpaired elderly

**DOI:** 10.1101/2021.01.22.21250293

**Authors:** Nicholas C. Cullen, Antoine Leuzy, Shorena Janelidze, Sebastian Palmqvist, Anna L. Svenningsson, Erik Stomrud, Jeffrey L. Dage, Niklas Mattsson-Carlgren, Oskar Hansson

## Abstract

Plasma biomarkers of amyloid, tau, and neurodegeneration (ATN) need to be characterized in cognitively unimpaired (CU) elderly indviduals. We therefore tested if plasma measurements of amyloid-β (Aβ)42/40, phospho-tau217 (P-tau217), and neurofilament light (NfL) together predict clinical deterioration in 435 CU individuals followed for an average of 4.8 ±1.7 years in the BioFINDER study. A combination of all three plasma biomarkers and basic demographics best predicted change in the cognition (Pre-Alzheimer’s Clinical Composite; R^2^=0.14, 95% CI [0.12-0.17]; P<0.0001) and subsequent AD dementia (AUC=0.82, 95% CI [0.77-0.91], P<0.0001). In a simulated clinical trial, a screening algorithm combining all three plasma biomarkers would reduce the required sample size by 70% (95% CI [54-81]; P<0.001) with cognition as trial endpoint, and by 63% (95% CI [53-70], P<0.001) with subsequent AD dementia as trial endpoint. Plasma ATN biomarkers show usefulness in cognitively unimpaired populations and could make large clinical trials more feasible and cost-effective.

## Introduction

Alzheimer’s disease (AD) is characterized by the presence of amyloid-β (Aβ) plaques and tau tangles.^1^ Specifically, Aβ is thought to potentiate the spread of neocortical tau pathology which in turn drives neurodegeneration and cognitive decline.^2^ Accruing evidence indicates, however, that these changes are slow and protracted and precede the initial symptoms of AD by many years or decades.^3,40^ A prerequisite for the testing of therapeutic interventions is the identification of individuals at risk for progression to AD dementia. Biomarkers have been explored for this.^41^ The National Institute on Aging and Alzheimer’s Association has proposed a framework for research wherein cerebrospinal fluid (CSF) and imaging (magnetic resonance and positron emission tomography (PET)) based measures of Aβ (A), tau (T), and neurodegeneration (N) can be compiled into an ATN classification system.^9^ CSF measures include the Aβ42/Aβ40 ratio, tau phosphorylated at threonine 181 (P-tau181) and neurofilament light (NfL). Development of new assays now makes it possible to measure ATN biomarkers in blood, including the Aβ42/Aβ40 ratio,^11,12^ P-tau^13,14,42^ and NfL.^15,16^ With lower cost and higher accessibility, blood-based AD biomarkers may circumvent the limitations inherent to CSF and imaging. While recent work on plasma biomarkers has primarily focused on individualized risk assessment in patients with mild cognitive impairment (MCI),^4-6^ it has been noted that substantial irreversible neuronal loss is already seen by this stage, which may reduce the likelihood of disease-modifying therapies to prevent dementia onset.^7^ This has led to an increasing focus on cognitively unimpaired (CU) older individuals at risk for progression to AD dementia on the basis of biomarker evidence of brain AD pathology.^8^ Studies on combinations of blood-based Aβ42/Aβ40, P-tau and NfL have reported findings on their diagnostic accuracy for the separation of AD dementia from CU individuals and patients with non-AD disorders,^11,13,17-21^ but data is lacking on their performance in predicting cognitive decline and clinical progression in CU individuals. We therefore tested the usefulness of plasma Aβ42/Aβ40, P-tau217, and NfL (separately and combined) for predicting cognitive decline and clinical outcomes in CU individuals who were followed longitudinally. We hypothesized that a combinations of ATN plasma biomarkers could improve the prediction of cognitive decline and risk of AD dementia compared to using only basic demographic information. We also investigated the extent to which plasma biomarkers could reduce the sample size required to adequately run AD clinical trials in a CU elderly population.

## Results

### Study population characteristics

A total of 435 CU individuals were included of which 167 (38.4%) had subjective cognitive decline (SCD). During follow-up, a total of 28 individuals converted to AD dementia (6.4%) and 39 individuals converted to all-cause dementia (9.0%). Participant characteristics are provided in Table 1. When testing the associations between unadjusted plasma biomarker values using Pearson’s correlation (Figure S1), there was a significant correlation between Aβ42/Aβ40 and P-tau217 (r=-0.22, P<0.0001), between P-tau217 and NfL (r=0.20, P=0.0005), and between NfL and Aβ42/Aβ40 (r=-0.10, P=0.03).

**Table 1.** Study participant characteristics.

|  | **Overall** | **CU** | **SCD** |
| --- | --- | --- | --- |
| **n** | 435 | 268 | 167 |
| **Age** | 72.58 (5.45) | 73.62 (5.01) | 70.92 (5.73) |
| **Education** | 12.17 (3.66) | 12.12 (3.74) | 12.27 (3.53) |
| **Sex = Male (%)** | 242 (55.6) | 159 (59.3) | 83 (49.7) |
| **MMSE, baseline** | 28.80 (1.21) | 29.03 (0.93) | 28.44 (1.47) |
| **MMSE, four-year change** | -1.02 (2.93) | -0.65 (1.79) | -1.68 (4.18) |
| **PACC, baseline** | 0.01 (0.74) | 0.17 (0.70) | -0.23 (0.75) |
| **PACC, four-year change** | -0.33 (0.85) | -0.27 (0.73) | -0.44 (1.05) |
| **Follow-up time** | 4.75 (1.66) | 4.99 (1.58) | 4.35 (1.71) |
| **Converted to AD dementia (%)** | 28 (6.4) | 3 (1.1) | 25 (15.0) |
| **Converted to any dementia (%)** | 39 (9.0) | 6 (2.2) | 33 (19.8) |
| **Plasma Aβ42/Aβ40, baseline** | 66.08 (7.28) | 66.26 (7.47) | 65.77 (6.97) |
| **Plasma P-tau217, baseline** | 4.97 (0.81) | 5.00 (0.77) | 4.92 (0.88) |
| **Plasma NfL, baseline** | 7.59 (0.46) | 7.63 (0.45) | 7.54 (0.47) |
| **CSF Aβ42/Aβ40, baseline** | 7.13 (0.50) | 7.15 (0.48) | 7.09 (0.53) |
| **CSF P-tau181, baseline** | 2.96 (0.40) | 2.94 (0.36) | 3.00 (0.45) |
| **CSF NfL, baseline** | 6.79 (0.45) | 6.75 (0.42) | 6.86 (0.50) |

This table shows the characteristics for the study population. All variables are presented as
“mean (standard deviation)” unless otherwise noted as a percentage “(%)” in the table. All
biomarkers besides plasma Aβ42/Aβ40 were natural log-transformed prior to analysis and all
biomarker values are provided in terms of pg/mL. Abbreviations: CU = cognitively
unimpaired; SCD = subjective cognitive decline; Aβ42/Aβ40 = the 42- divided by the 40-
amino acid long beta-amyloid peptide; P-tau217 (P-tau181): the (181-) 217-amino acid long
phosphorylated tau peptide; NfL: neurofilament light chain

A total of 201 participants (46.2%) were assigned as plasma Aβ42/Aβ40-positive, 162 participants (37.2%) as plasma P-tau217-positive, and 165 participants (37.9%) as plasma NfL-positive.

### Modelling longitudinal change in cognition

We first tested whether plasma biomarkers were significantly associated with cognitive decline as measured by the Pre-Alzheimer’s Clinical Composite (PACC). When added individually to a basic model consisting of age, sex, and education (Figure 1), we found that plasma P-tau217 had the strongest association with longitudinal change in PACC (β=-0.20 points per year per SD increase in biomarker value, P<0.0001), followed by plasma Aβ42/Aβ40 (β=-0.18, P=0.0002), and plasma NfL (β=-0.16, P=0.001). When combining all three plasma biomarkers together in the same model (Table 2), all three biomarkers remained significant (P=0.004 for Aβ42/Aβ40, P=0.002 for P-tau217, P =0.01 for NfL). Adding *APOE* ε4 status to the basic model did not affect the significance of any biomarkers in either the combined or individual models (Table S1), although the effect of plasma Aβ42/Aβ40 in the combined model decreased the most (β=-0.16, P=0.002 with *APOE* ε4 status).

**Table 2.** Association between plasma biomarkers and longitudinal PACC.

| Model | Beta Coefficient |  |  | R<sup>2</sup><br>[95% CI] | Ref: Basic Model |  |
| --- | --- | --- | --- | --- | --- | --- |
| | Plasma<br>Aβ42/Aβ40 | Plasma<br>P-tau217 | Plasma<br>NfL | | P value | AIC<sub>Δ</sub> |
| ATN | -0.15<br>(P=0.0026) | -0.15<br>(P=0.0020) | -0.12<br>(P=0.0141) | 0.14<br>[0.12, 0.17] | <0.0001 | -28 |
| A | -0.19<br>(P=0.0001) |  |  | 0.11<br>[0.09, 0.14] | <0.0001 | -14 |
| T |  | -0.20<br>(P<0.0001) |  | 0.09<br>[0.08, 0.13] | 0.0002 | -13 |
| N |  |  | -0.15<br>(P=0.0016) | 0.10<br>[0.08, 0.14] | 0.0015 | -9 |

This table shows the results from fitting linear mixed effects models with longitudinal PACC as outcome and plasma biomarkers added separately or all together to a basic model consisting of age, sex, and education. Beta coefficients are presented in terms of “PACC points / year per standard deviation change in biomarker value.” R<sup>2</sup> values were evaluated and confidence intervals were calculated using 1000 bootstrapped samples. The basic model consisting of only demographics had R<sup>2</sup> = 0.07 (95% CI [0.06, 0.11]) and AIC = 6699.
Legend: P-values represent an ANOVA comparison to the basic model; AIC<sub>Δ</sub> values represent the change in AIC compared to the basic model and an AIC<sub>Δ</sub> value of -2 or lower implies a better fit than the basic model.

**Figure 1.**
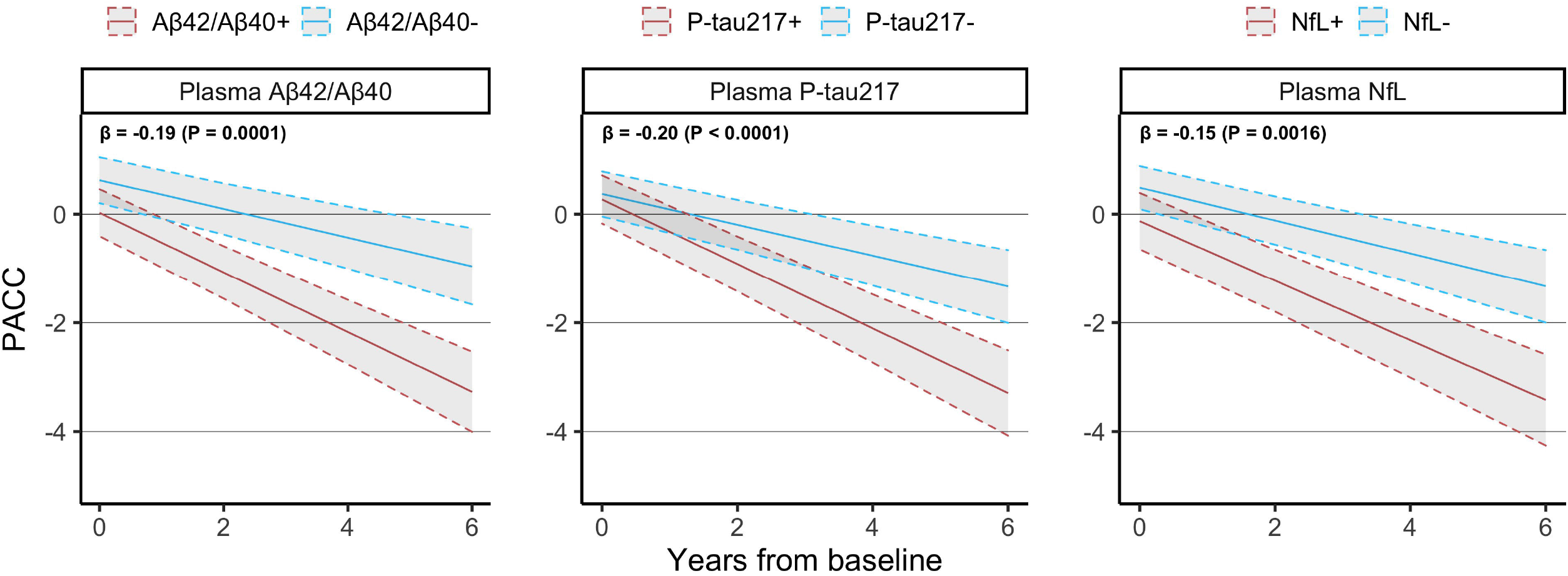
Relationship between plasma biomarkers and change in PACC. This figure shows the longitudinal PACC trajectory estimated for a CU individual with average age, average education, female sex and either biomarker-negative or biomarker-positive. Beta coefficients at the top left of each panel are presented in terms of “points / year per standard deviation change in biomarker value” and are derived from linear mixed effects models with longitudinal PACC as outcome and age, sex, education, plus each plasma biomarker included separately from each other.

All plasma biomarker models significantly improved prediction of PACC change compared to the basic model (AIC=6699; R^2^ = 0.07, CI [0.06, 0.11]; see Table 2). In terms of AIC, a combination of all three plasma biomarkers predicted PACC change best (AIC_Δ_=-28 versus basic model; R^2^=0.14 [0.12, 0.17], P<0.0001), followed by plasma Aβ42/Aβ40 only (AIC_Δ_=-14; R^2^=0.11 [0.09, 0.14], P<0.0001), plasma P-tau217 only (AIC_Δ_ =-13; R^2^=0.09 [0.08, 0.13], P=0.0002), and finally plasma NfL only (AIC_Δ_=-9; R^2^=0.10 [0.08, 0.14], P=0.002).

A sensitivity analysis in which longitudinal MMSE was used as the cognitive outcome showed an increased effect of plasma NfL and a decreased effect of plasma Aβ42/Aβ40, while plasma P-tau217 remained a strong predictor (Figure S2; Table S2). All biomarkers remained significant when combined together. A comparison of model results between corresponding CSF and plasma biomarkers indicated that – in terms of AIC – CSF biomarkers had significantly better combined prediction of PACC change than plasma biomarkers (AIC_ΔCSF_ = -98 versus AIC_ΔPlasma_ = -28; Table S3).

### Modelling clinical progression to dementia

We next tested whether plasma biomarkers were significantly associated with subsequent development of dementia. When added individually to a basic model consisting of age, sex, and education (Figure 2), we found that plasma P-tau217 had the strongest association with conversion to AD dementia (HR=3.54 increased odds / std., P<0.0001), followed by plasma Aβ42/Aβ40 (HR=2.00, P=0.0002), while plasma NfL on its own did not add significant information to the basic model (HR=1.51, P=0.07). When combining all three plasma biomarkers together in the same model (Table 3), plasma P-tau217 (HR=2.97, P=0.0004) and Aβ42/Aβ40 (HR=1.83, P=0.003) remained significant. When adding *APOE* ε4 status to the set of demographic predictors, AUC values increased for all models, but no change in significance for any plasma biomarkers was observed (Table S4).

**Table 3.** Association between plasma biomarkers and conversion to AD dementia.

| **Model** | **Hazard Ratio** |  |  | **AUC**<br>**[95% CI]** | **Ref: Basic Model** |  |
| --- | --- | --- | --- | --- | --- | --- |
|  | **Plasma**<br>**Aβ42/Aβ40** | **Plasma**<br>**P-tau217** | **Plasma**<br>**NfL** |  | **P value** | **AIC$_{\Delta}$** |
| ATN | 1.83<br>(P=0.0031) | 2.97<br>(P=0.0004) | 1.07<br>(P=0.7898) | 0.82<br>[0.77, 0.91] | <0.0001 | -25 |
| A | 2.00<br>(P=0.0002) |  |  | 0.77<br>[0.70, 0.86] | 0.0003 | -11 |
| T |  | 3.54<br>(P<0.0001) |  | 0.77<br>[0.68, 0.86] | <0.0001 | -21 |
| N |  |  | 1.51<br>(P=0.0719) | 0.67<br>[0.60, 0.78] | 0.0753 | -1 |

This table shows the results from fitting Cox regression models with conversion to AD as outcome and plasma biomarkers added separately or all together to a basic model consisting of age, sex, and education. Hazard ratios are presented in terms of “increased risk of converting to AD for each standard deviation change in biomarker value.” AUC values were evaluated and confidence intervals were calculated using 1000 bootstrapped samples. The basic model consisting of only demographics had AUC = 0.64 (95% CI [0.55, 0.77]) and AIC = 274. Legend: P-values represent an ANOVA comparison to the basic model; AIC $_{\Delta}$ values represent the change in AIC compared to the basic model and an AIC $_{\Delta}$ value of -2 or lower implies a better fit than the basic model

**Figure 2.**
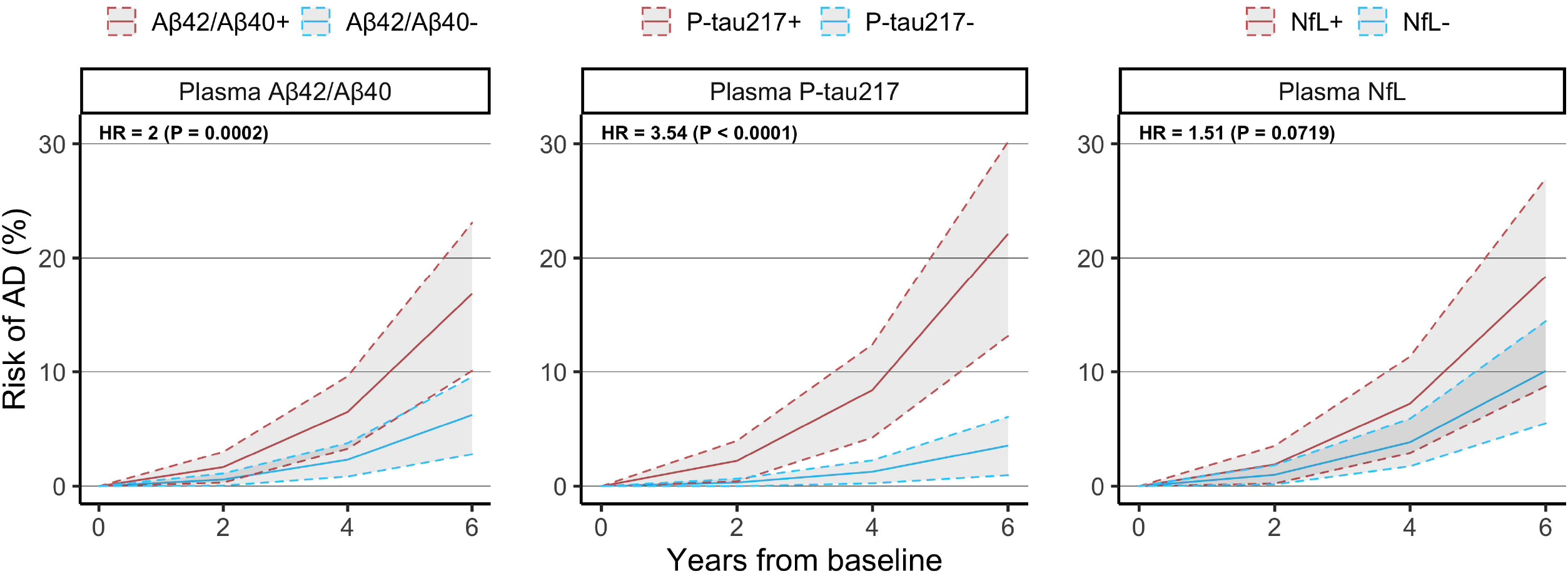
Relationship between plasma biomarkers and conversion to AD. This figure shows the conversion to AD dementia estimated for a CU individual with average age, average education, female sex and either biomarker-negative or biomarker-positive. Hazard ratios at the top left of each panel are presented in terms of “increased risk of converting to AD dementia per standard deviation change in biomarker value” and are derived from Cox regression models with conversion to AD dementia as outcome and age, sex, education, plus each plasma biomarker included separately from each other.

All plasma biomarker-based models significantly improved prediction of four-year conversion to AD compared to the basic model (AIC=274; AUC=0.64 [0.55, 0.77]; see Table 3). In terms of AIC, the model with all three plasma biomarkers together fit the data best (AIC_Δ_=-25 versus basic model; AUC=0.82 [0.77, 0.91], P<0.0001), followed by plasma P-tau217 only (AIC_Δ_=-31; AUC=0.77 [0.68, 0.86], P<0.0001), plasma Aβ42/Aβ40 only (AIC_Δ_=-11; AUC=0.77 [0.70, 0.86], P= 0.0003). The model with plasma NfL alone was not significantly better than demographics only (AIC_Δ_=-1; AUC=0.67 [0.60, 0.78], P=0.08).

A sensitivity analysis in which conversion to all-cause dementia was used as the outcome of interest produced overall similar results, but with plasma NfL now adding significant prognostic information compared to when predicting conversion to AD dementia (Figure S3; Table S5).

### Power analysis for theoretical clinical trial

Finally, we tested whether using plasma biomarkers for participant screening could reduce the required sample size of a theoretical clinical trial aimed at slowing change in PACC by 30% over four years compared to a similar clinical trial without any biomarker-based inclusion screening (Figure 3; Table S6). Using pre-defined cutoffs for the inclusion threshold (see Methods S2 for cutoff values), we found that a significantly lower sample size was required when enriching for plasma P-tau217 (sample size reduction [SS_Δ_]=47%, CI [16, 65], P=0.007), plasma Aβ42/Aβ40 (SS_Δ_=45%, CI [20, 63], P=0.003) and plasma NfL (SS_Δ_=41%, CI [5, 63], P=0.03). Moreover, combining all three biomarkers in a multivariable enrichment model led to the largest reduction in sample size (SS_Δ_=70% [54, 81], P<0.001). All plasma biomarkers were robust to up to 20% change in inclusion threshold and a stricter inclusion threshold led to larger reduction in required sample size (Figure 4).

**Figure 3.**
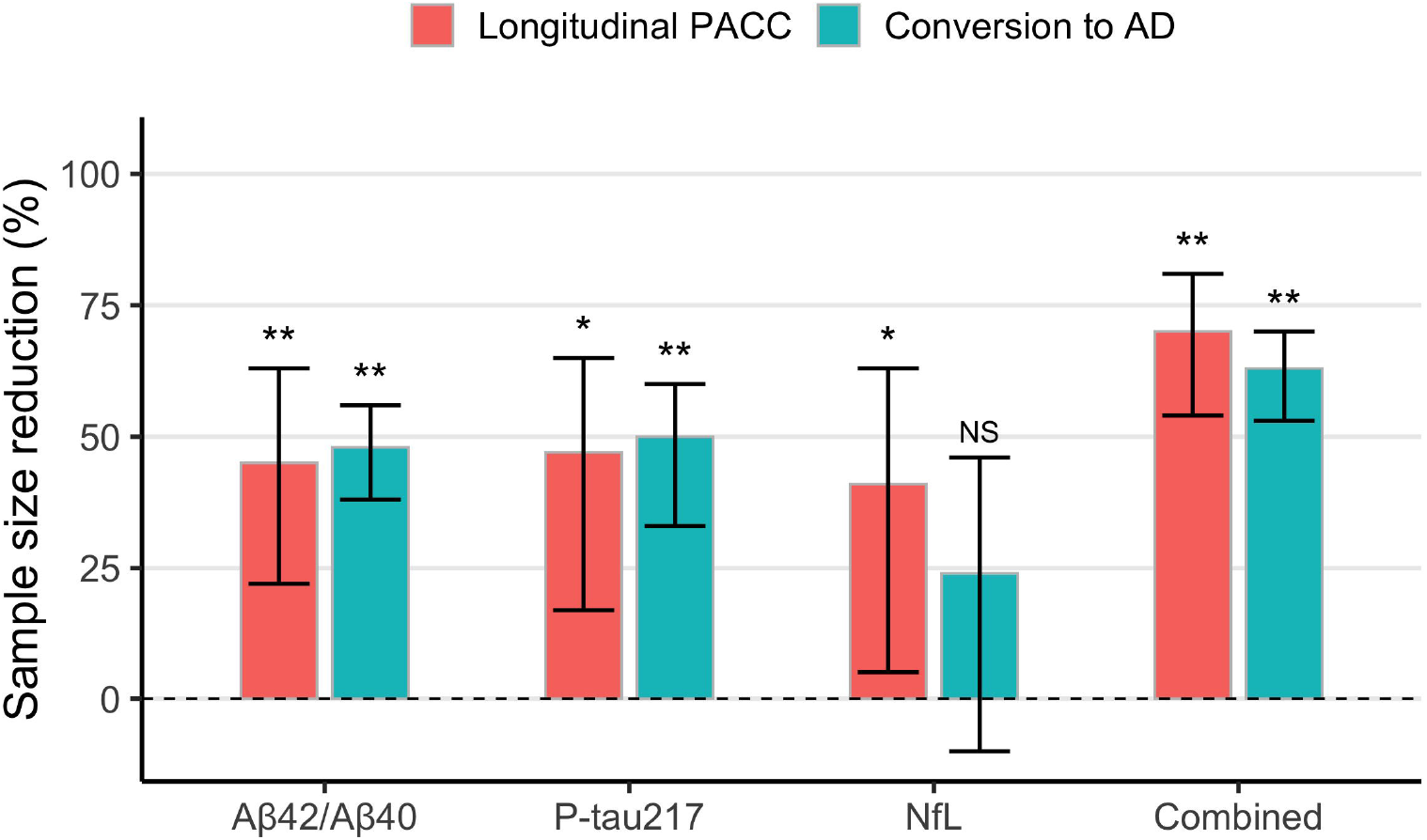
Power enrichment analysis of plasma biomarkers in a theoretical clinical trial. This figure shows the reduction in sample size resulting from using plasma biomarkers for inclusion enrichment in theoretical clinical trials aimed at slowing decline in PACC or reducing risk of conversion to AD dementia in a CU population. Sample sizes were estimated for a trial enriched using pre-defined cutoffs for each biomarker as inclusion threshold. Confidence intervals were derived using 1000 bootstrapped trials. Legend: *=P<0.05, **=P<0.005, NS=Not Significant.

**Figure 4.**
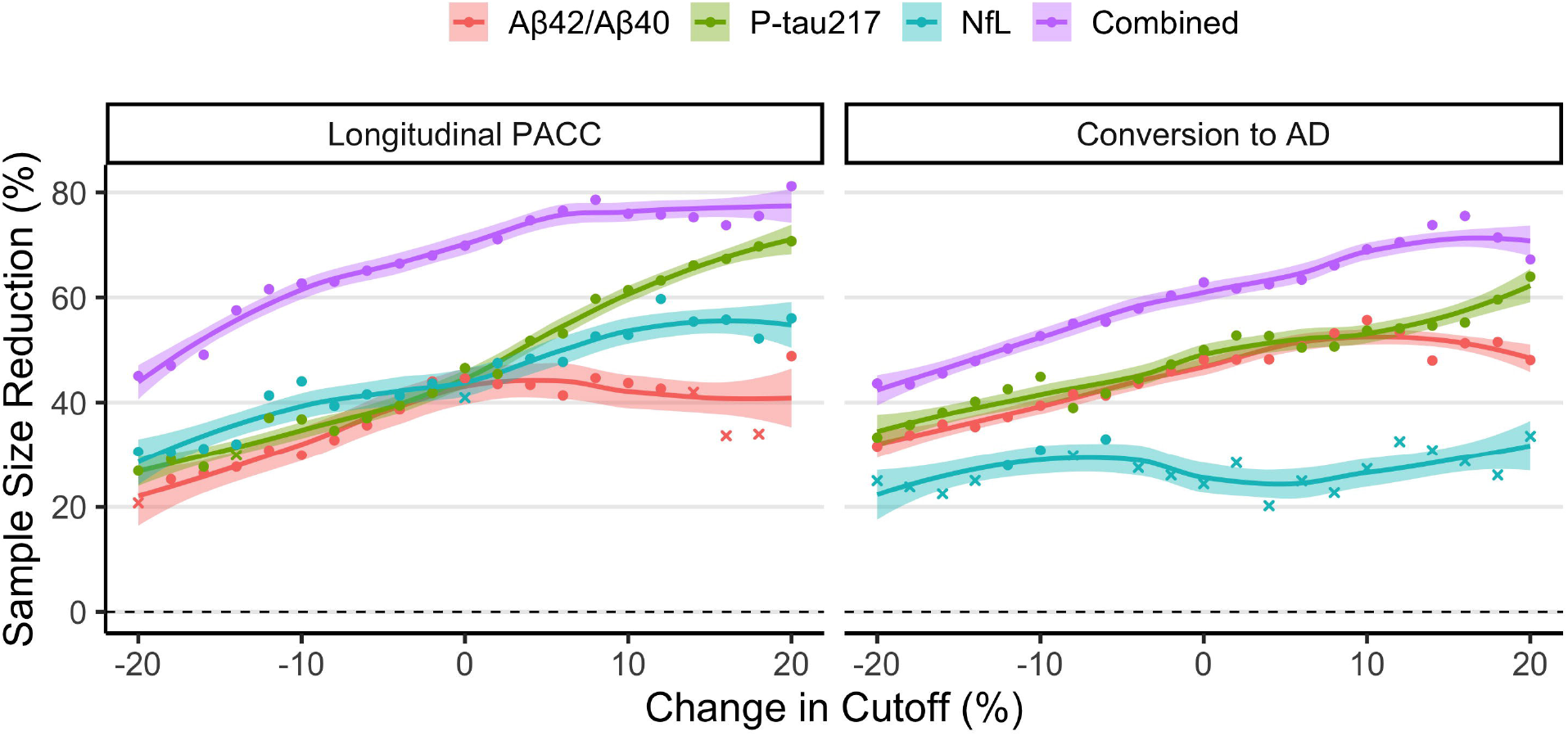
Effect of inclusion threshold variability on biomarker effectiveness. This figure shows the effect that varying the inclusion threshold by 20% in either direction of the pre-defined cutoff has on the reduction in sample size in a theoretical clinical trial using biomarkers for inclusion enrichment. The reduction in sample size is compared to an unenriched trial with PACC or conversion to AD dementia as primary outcome. For datapoints in the figure, “circles” represent cutoffs for which required sample size was significantly reduced, while “crosses” represent cutoffs for which required sample size was not significantly reduced.

For a clinical trial aimed at reducing the risk of conversion to AD by 30% over four years (Figure 3; Table S6), a significant reduction in required sample size was again observed when enriching for plasma P-tau217 (SS_Δ_=48%, CI [38, 56], P<0.001) and plasma Aβ42/Aβ40 (SS_Δ_=48%, CI [35, 60], P=0.001), while plasma NfL did not significantly enrich for such a clinical trial (SS_Δ_=24%, CI [-10, 45], P=0.20). An enrichment model which combined all three biomarkers again achieved greatest reduction in sample size (SS_Δ_=63%, CI [53, 70], P<0.001). Again, all plasma biomarkers besides plasma NfL appeared robust to larger variation in the inclusion threshold (Figure 4).

## Discussion

We tested the hypothesis that plasma biomarkers (Aβ42/Aβ40, P-tau217 and NfL) previously established in AD research can improve prediction of cognitive and clinical outcomes for elderly CU individuals. Our primary findings were that plasma biomarkers add significant prognostic information to a basic model consisting only of demographic information (age, sex and education) and can significantly reduce the number of participants required to run AD-related clinical trials in this population.

All three plasma biomarkers were significantly associated with the primary outcomes on their own, but we showed here that they also provide some degree of overlapping information with each other. Plasma P-tau217 and Aβ42/Aβ40 contributed significantly to the prediction of both PACC and risk of AD dementia, while plasma NfL added significant prognostic information for PACC and appeared more useful for predicting general cognitive decline (MMSE) and all-cause dementia. Together, our results demonstrate the wide applicability of this plasma biomarker panel in a CU elderly population.

All three plasma biomarkers also significantly reduced sample sizes needed to run AD clinical trials in a CU population when used for pre-trial inclusion screening. This is a promising result given that clinical trials of AD are increasingly focused on elderly individuals without existing cognitive impairment. Running such a clinical trial in a cost-effective way likely requires the ability to easily and inexpensively identify CU individuals at risk for developing AD. Our results suggest that these plasma biomarkers are in fact an effective way to enrich clinical trials in a cognitively normal population. While the biomarkers showed promise for stand-alone use for trial enrichment, combining these biomarkers in a multivariable model led to the greatest reduction in required trial sample size for PACC and the lowest variability in estimated sample size reduction.

Interestingly, the combined multivariate model and individual plasma biomarkers were robust to variation in the inclusion threshold, indicating that random shifts in the demographics of a clinical trial screening population or in the analytical variability of the plasma assay may not have a significant negative impact on enrichment. Still, this result should be interpreted with caution since it is not easy to determine how much of the variability is controllable (e.g. through pre-analytical factors) and how much is due to inherent biological variability. It was also not possible to account for the possibility that enrichment may increase study attrition rates or otherwise alter trial dynamics since this type of data is not available to us.

With regards to cognitive outcomes, we found that all three plasma biomarkers added independent information for predicting change in PACC even when combined in the same model. This indicates that if only one or two of the three biomarkers have already been collected in a specific individual, a more accurate prediction can be obtained by additionally measuring the other, unkown biomarker(s). This result was not dependent on the inclusion of *APOE* ε4 status, although including such information did generally raise the prognostic accuracy. Also, plasma Aβ42/Aβ40 had a stronger effect with PACC as outcome rather than MMSE; this might be expected given that PACC was created specifically to pick up early AD-related changes in non-demented individuals^23^ and has been shown to have greater sensitivity to Aβ-related cognitive decline,^*28*^ while the MMSE is a measure of global cognition originally created to differentiate dementia patients from those with psychiatric syndromes.^31,32^ This highlights the idea that selection of cognitive scale impacts which plasma biomarkers are most relevant.

With regards to clinical outcomes, we found that both plasma Aβ42/Aβ40 and plasma P-tau217 added independent information for prediction of conversion to AD dementia, even when combined in the same model, further adding to the existing evidence of plasma Aβ42/Aβ40 and P-tau217 as AD-specific markers.^29,30^ The contribution of plasma Aβ42/Aβ40 was greatly affected when *APOE* ε4 status was included, but the same effect was not seen for CSF Aβ42/Aβ40, indicating that *APOE* ε4 status may contain prognostic information related to Aβ accumulation which is not picked up by plasma Aβ42/Aβ40.^33^ This finding aligns with studies showing a link between *APOE* ε4 and alterations in Aβ-related processes.^34,35^ Our results further suggest that *APOE* ε4 status may be informative in a CU population even when plasma biomarker values are available, as prediction of subsequent dementia in particular was significantly better when including *APOE* ε4 as a covariate. The effect of plasma NfL greatly increased when analyzing conversion to all-cause dementia, thereby strengthening previous findings of plasma NfL as a general biomarker of neurodegeneration that detects non-AD related neurodegenerative changes. ^36-38^

We also compared plasma biomarkers directly against their corresponding CSF biomarkers. CSF biomarkers still appear to be superior to plasma biomarkers in terms of cognitive and clinical prognosis in elderly individuals without cognitive impairment, although this comparison was based on AIC values and there was in fact significant overlap in R^2^ between models. This indicates that CSF and plasma may provide similar predictive accuracy overall but that CSF provides a much more confident prediction. We hypothesize that this difference is largely based on the observation that CSF Aβ42/Aβ40 is a much stronger prognostic marker than plasma Aβ42/Aβ40. However, the descrepency between plasma and CSF Aβ42/Aβ40 may not be biological but may instead be related to the performance of the specific plasma assays used here (Elecsys immunoassay). Our work in MCI patients showed a greater effect of plasma Aβ42/Aβ40 by using a mass-spectrometry method, indicating that a more effective plasma assay could lead to greater value.^39^ However, such data was not widely available in this cohort. Further work is needed to compare performance of CSF and plasma biomarkers in CU individuals using different assays, as recent studies suggest that the difference between CSF and plasma biomarkers is less pronounced in MCI individuals.^39^

The strengths of this study include the standardized collection and measurement of relevant biomarkers in both plasma and CSF—which allowed for direct comparison between biomarkers and across the two modalities—and the availability of multi-year follow-up data in a large number of participants. Our study improves in particular on previous studies linking novel plasma tau levels on their own with cognitive decline and neurodegeneration in similar populations^43,44^. Understanding the overlapping contributions of novel tau biomarkers is therefore a major contribution of our results. Not having a state-of-the-art mass-spectrometry plasma Aβ42/Aβ40 assay is a weakness, although plasma Aβ42/Aβ40 was still important to the prognostic algorithms. Additionally, since evidence of abnormal CSF biomarker levels was required for setting the diagnosis of AD dementia at the memory clinic, it was not possible to fairly compare the performance of plasma and CSF biomarkers in this manner. The evidence we did provide for cognition, however, indicates that CSF biomarkers are still superior to plasma biomarkers in a CU elderly population at least in terms of pure prediction. Factoring in cost and ease of collection may make plasma more beneficial at a more general level. In the future, we hope to validate our findings in an independent cohort as soon as other studies with sufficient longitudinal cognitive follow-up and measurements of plasma P-tau217 (a relatively new biomarker) become available.

To summarize, our results show that plasma biomarkers relate to AD-related changes in the CU elderly and can significantly reduce sample sizes needed to run clinical trials in such a population. Further work is needed to validate the optimal panel of biomarkers in an eldery CU population, although our preliminary results suggest that all three core plasma biomarkers could be useful.

## Methods

### Participants

The Swedish BioFINDER (Biomarkers for Identifying Neurodegenerative Disorders Early and Reliably; clinical trial no. NCT01208675, www.biofinder.se) cohort used in the present analysis consisted of CU individuals (i.e. no objective evidence of cognitive impairment at baseline) and SCD individuals (i.e. individuals who were referred to the memory clinic for investigation but deemed to not have cognitive impairment) with available plasma Aβ42/Aβ40, P-tau217, and NfL measurements and CSF Aβ42/Aβ40, P-tau181, and NfL measurements. Inclusion and exclusion criteria have been described elsewhere^22^ and are included in the Supplement (eMethods 1). Longitudinal follow-up visits occurred every two years for CU individuals and every year for SCD individuals. Conversion to a clinical diagnosis of dementia (AD dementia or dementia due to any cause [“all-cause dementia”]) during follow-up was evaluated according to the diagnostic and statistical manual of mental disorders version 5 (DSM-5) criteria and required a positive indication on CSF-based AD biomarkers. All participants gave written informed consent. Ethical approval was given by the Regional Ethical Committee in Lund, Sweden.

### Cognitive and clinical outcomes

The primary cognitive outcome was the Preclinical Alzheimer’s Cognitive Composite (PACC), as this scale was developed specifically to identify early cognitive changes in individuals without dementia.^23^ The PACC score used for the present study was comprised of the MMSE, delayed word recall from the Alzheimer’s Disease Assessment Scale (ADAS)-Cognitive Subscale (weighted double to reflect the emphasis on memory tests in the original PACC), animal fluency, and trail-making B tests and was calculated as described previously.^23,24^ The secondary cognitive outcome was the Mini-Mental State Examination (MMSE) collected longitudinally during all available follow-up visits. The primary clinical outcome was conversion to AD dementia at any time during longitudinal follow-up. The secondary clinical outcome was conversion to all-cause dementia.

### Plasma and CSF biomarker assays

Biomarkers included in the present study were Aβ42/Aβ40, P-tau217, and NfL measured in plasma, along with Aβ42/Aβ40, P-tau181 (P-tau217 was not available in CSF), and NfL measured in CSF. Plasma Aβ42/Aβ40 was measured using an Elecsys immunoassay on a Cobas e601 analyzer (Roche Diagnostics GmbH, Penzberg, Germany).^11^ Plasma P-tau217 was measured on a Meso-Scale Discovery platform (MSD, Rockville, MD), using an assay developed by Eli Lilly.^10^ Plasma NfL was analyzed using a Simoa-based assay.^16^ CSF levels of Aβ42/Aβ40 and P-tau181 were measured using Elecsys assays (Roche Diagnostics GmbH), and CSF NfL was measured using the ELISA method (UmanDiagnostics AB, Umeå, Sweden). All biomarker values were transformed using the natural log function and any biomarker values more than four standard deviations away from the mean were excluded.

Biomarker cutoffs were derived using the Youden index procedure to maximize separation of CU individuals without a positive CSF Aβ42/Aβ40 (n=350) who did not convert to AD dementia versus MCI (n=111) and CU (n=30) individuals who did convert to AD dementia).

### Statistical Analysis

Linear mixed effects (LME) modelling was used to model longitudinal PACC, with five models tested in total: a basic model (age, sex, education), the basic model plus each of the three plasma biomarkers added separately (i.e. basic model + Aβ42/Aβ40; basic model + P-tau217; basic model + NfL), and the basic model plus all three plasma biomarkers together. LME models had random intercepts and random slopes with an unstructured covariance matrix. Cox regression was used to model clinical conversion to AD dementia with the same models fit as above; all participants were right censored at last follow-up visit or at clinical conversion. A power analysis was performed in which the reduction in sample size needed to achieve 80% power to observe a 30% reduction in cognitive decline or clinical conversion was calculated in a scenario where each biomarker was used individually (based on univariate thresholds) or combined (based on a fitted multivariate model) to screen individuals for trial inclusion and in order to enrich the trial population for the relevant outcome.

The primary cognitive outcome was also analyzed using CSF biomarkers on the same subjects and results were compared between CSF and plasma models using the Akaike Information Criterion (AIC). The larger the difference in AIC is between two models, the less plausible it is that the model with higher AIC provides the best fit.^25,26^ Risk of AD dementia was not analyzed using CSF biomarkers because these biomarkers were consulted by neurologists during diagnosis, leading to potential circular reasoning.

Sensitivity analyses were also performed with (a) MMSE as the cognitive outcome instead of PACC, (b) all-cause dementia as the clinical outcome instead of AD dementia, and (c) *APOE* ε4 status (one or more ε4 copy versus zero ε4 copies) included as an additional predictor in the basic model.

All analyses were performed using the *ABA* (“Automated Biomarker Analysis”) package (v0.0.1) written in the R programming language (v4.0.0). The *ABA* package was created specifically to ensure complete reproducibility of all results achieved here. All statistical tests were two-sided with a significance level of 0.05.

## Data Availability

For more information visit Biofinder.se. Relevant data from the study and code to perform the statistical analysis herein may be made available upon request from qualified researchers.

## Acknowledgements

We would like to acknowledge all of the BioFINDER team members as well as participants in the study and their family members for their dedication and patience. Work at the authors’ research center supported by the Swedish Research Council (2016-00906), the Knut and Alice Wallenberg foundation (2017-0383, and WCMM fellowship for Mattsson-Carlgren), the Medical Faculty at Lund University (WCMM fellowship for Mattsson-Carlgren), Region Skåne (WCMM fellowship for Mattsson-Carlgren), the Marianne and Marcus Wallenberg foundation (2015.0125), the Strategic Research Area MultiPark (Multidisciplinary Research in Parkinson’s disease) at Lund University, the Swedish Alzheimer Foundation (AF-745911, AF-930655), the Swedish Brain Foundation (FO2019-0326, FO2019-0029), The Parkinson foundation of Sweden (1280/20), the Skåne University Hospital Foundation (2020-O000028), Regionalt Forskningsstöd (2020-0314), the Swedish federal government under the ALF agreement (2018-Projekt0279), Stiftelsen Gamla Tjänarinnor (2019-00845), EU Joint Programme – Neurodegenerative Disease Research (2019-03401), The Bundy Academy, and The Konung Gustaf V:s och Drottning Victorias Frimurarestiftelse.

## Author Contributions

Concept and design: Cullen, Mattsson-Carlgren, Hansson

Data Acquisition: Stomrud, Svenningsson, Dage, Palmqvist, Mattsson-Carlgren, Hansson

Statistical analysis: Cullen, Leuzy, Svenningsson

Drafting of the manuscript: Cullen, Leuzy, Mattsson-Carlgren, Hansson

Critical revision of the manuscript for important intellectual content: All authors;

Obtained funding: Hansson, Mattsson-Carlgren, Palmqvist

Administrative, technical, or material support: Stomrud

Supervision: Mattsson-Carlgren, Hansson

## Materials & Correspondance

All requests and correspondence should be addressed to Niklas Mattsson-Carlgren, MD, PhD or Oskar Hansson, MD, PhD.

---

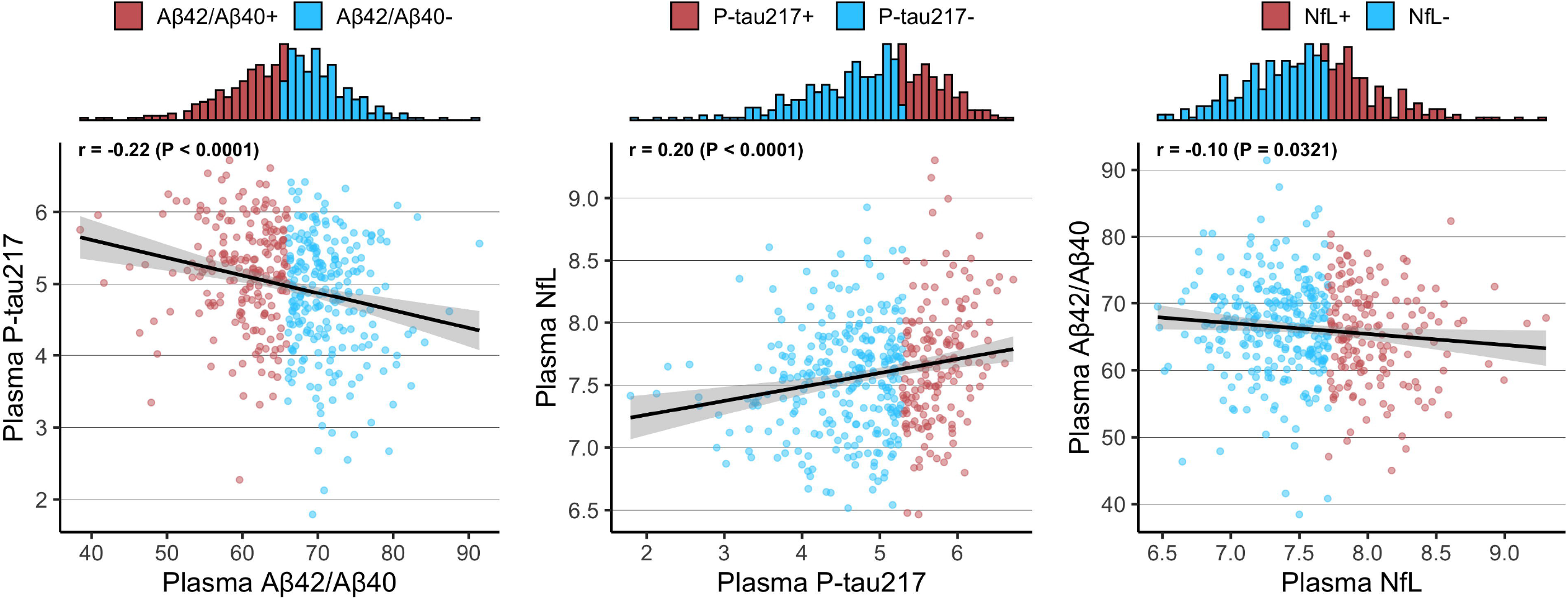

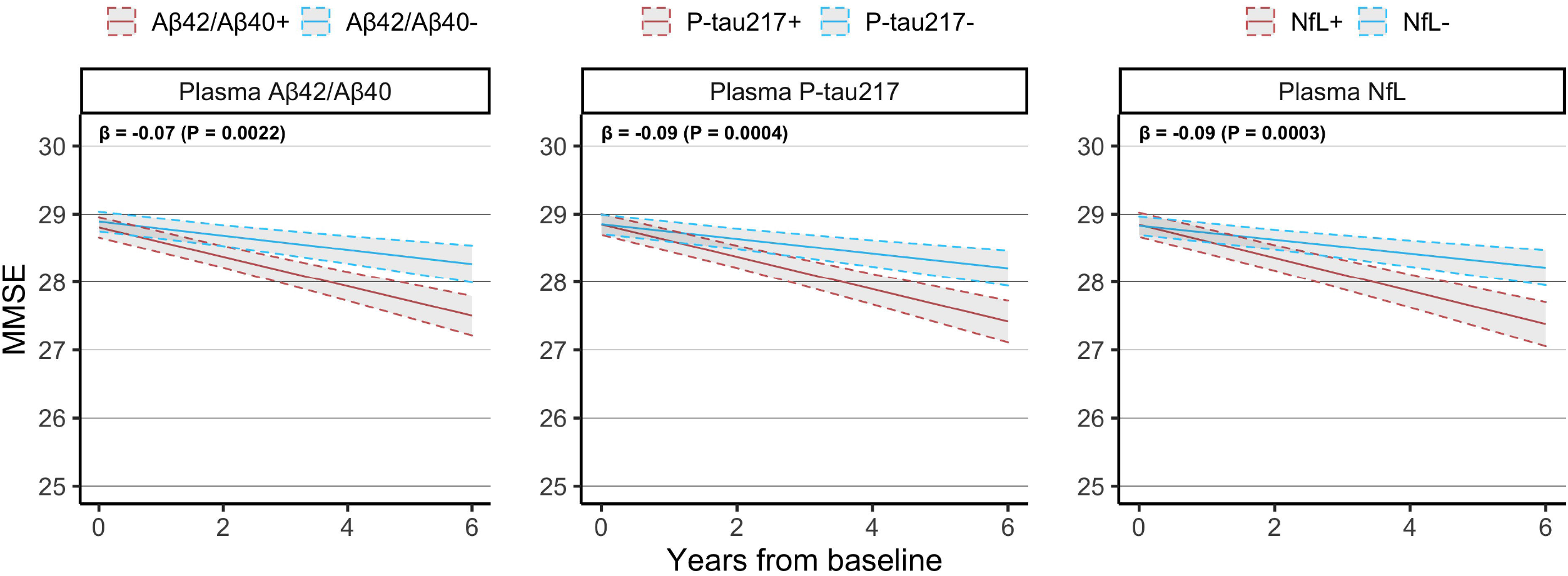

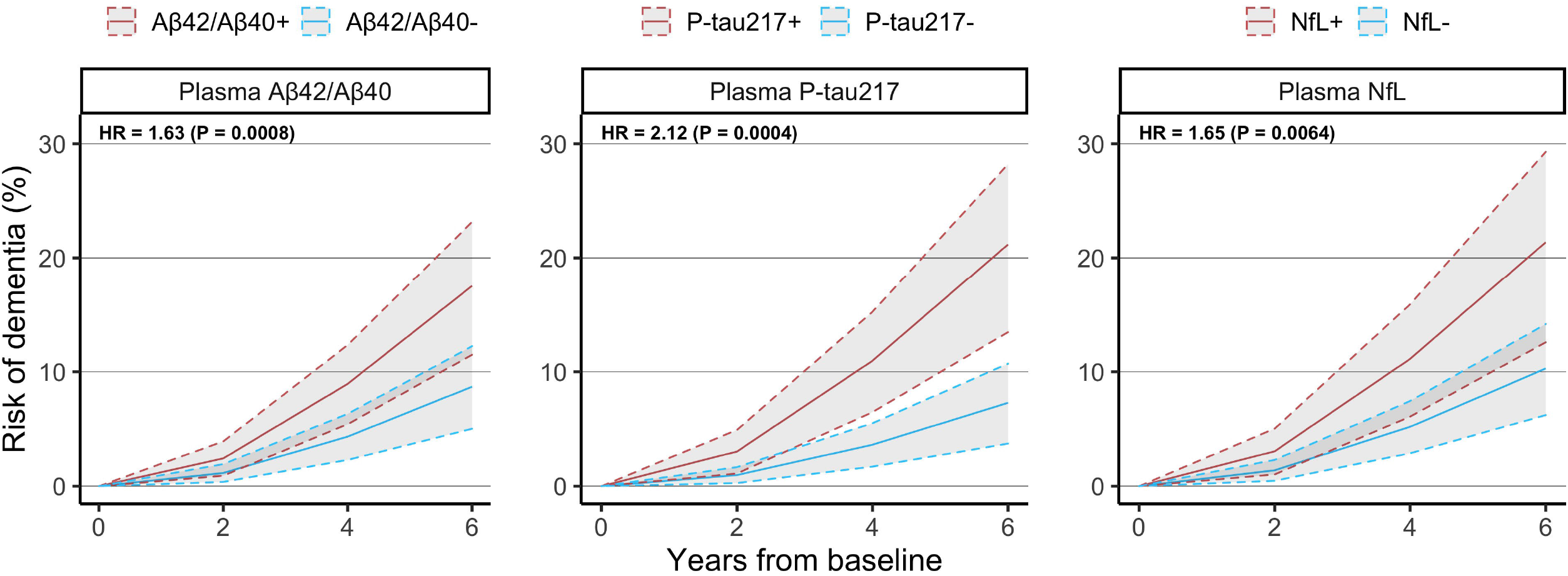

