## Supplementary Material for "Plasma biomarkers of Alzheimer’s disease predict cognitive decline and could improve clinical trials in the cognitively unimpaired elderly"

**Supplementary Materials**

**Plasma amyloid, phosphorylated tau, and neurofilament light predict Alzheimer-associated changes and could improve clinical trials in cognitively unimpaired elderly**

Nicholas C. Cullen, B.S.^1^, Antoine Leuzy, Ph.D.^1^, Shorena Janelidze, Ph.D.^1^, Sebastian Palmqvist, M.D.^1,2^, Anna L. Svenningsson, M.D.^1,2^, Erik Stomrud, M.D.^1,2^, Jeffrey L. Dage, Ph.D.^3^, Niklas Mattsson-Carlgren, M.D.^1,4,5, †^, Oskar Hansson, M.D.^1,2, †^

^1^Lund University, Clinical Memory Research Unit, Lund, Sweden; ^2^Memory Clinic, Skåne University Hospital, Lund, Sweden; ^3^Eli Lilly and Company, Indianapolis, IN, USA; ^4^Department of Neurology, Skåne University Hospital, Lund, Sweden; ^5^Wallenberg Centre for Molecular Medicine, Lund University, Lund, Sweden

^†^ Contributed equally as senior authors

**Supplementary Methods**

**Methods S1. Inclusion and exclusion criteria for CU participants in BioFINDER**

The inclusion criteria for CU individuals in BioFINDER are : i) age ≥65 years; ii) absence of cognitive symptoms as assessed by a physician with special interest in cognitive disorders; iii) MMSE score between 26 and 30 at screening visit; iv) do not fulfill the criteria for MCI or dementia according to DSM-5 (American Psychiatric Association, 2013); v) fluent in Swedish. Exclusion criteria include i) significant unstable systemic illness that makes it difficult to participate in the study; ii) current significant alcohol or substance misuse; iii) refusing lumbar puncture or MRI.

**Methods S2. Binary cutoffs for plasma biomarkers**

The following cutoffs were determined using the Youden index procedure to maximize separation of CU individuals without a positive CSF Aβ42/Aβ40 indication (n=350) who did not convert to AD dementia versus MCI (n=111) and CU (n=30) individuals who did convert to AD dementia:

- Plasma Aβ42/Aβ40: 0.066 pg/mL
- Plasma P-tau217: 0.199 pg/mL
- Plasma NfL: 22.3 pg/mL

**Supplementary Tables**

**Table S1. Association between plasma biomarkers and longitudinal PACC with APOE included in basic model**

| **Model** | **Beta Coefficient** | | | **R^2^**  **[95% CI]** | **Ref: Basic Model** | |
| --- | --- | --- | --- | --- | --- | --- |
|  | **Plasma**  **Aβ42/Aβ40** | **Plasma**  **P-tau217** | **Plasma**  **NfL** |  | **P value** | **AIC_Δ_** |
| ATN | -0.15 (P=0.0025) | -0.15 (P=0.0020) | -0.12 (P=0.0142) | 0.15  [0.12, 0.17] | <0.0001 | -28 |
| A | -0.19 (P=0.0001) |  |  | 0.11  [0.09, 0.14] | 0.0002 | -14 |
| T |  | -0.20 (P<0.0001) |  | 0.10  [0.08, 0.13] | 0.0001 | -14 |
| N |  |  | -0.15 (P=0.0017) | 0.10  [0.09, 0.14] | 0.0016 | -9 |

This table shows the results from fitting linear mixed effects models with longitudinal PACC as outcome and plasma biomarkers added separately or all together to a basic model consisting of age, sex, education, and *APOE* status. Beta coefficients are presented in terms of “PACC points / year per standard deviation change in biomarker value.” R^2^ values were evaluated at the four-year follow-up point, and confidence intervals were calculated using 1000 bootstrapped samples. The basic model consisting of only demographics had R^2^ = 0.07 (95% CI [0.07, 0.11]) and AIC = 6700. Legend: P-values represent an ANOVA comparison to the basic model; AIC_Δ_ values represent the change in AIC compared to the basic model and an AIC_Δ_ value of -2 or lower implies a better fit than the basic model.

**Table S2. Association between plasma biomarkers and longitudinal MMSE**

| **Model** | **Beta Coefficient** | | | **R^2^**  **[95% CI]** | **Ref: Basic Model** | |
| --- | --- | --- | --- | --- | --- | --- |
|  | **Plasma**  **Aβ42/Aβ40** | **Plasma**  **P-tau217** | **Plasma**  **NfL** |  | **P value** | **AIC_Δ_** |
| ATN | -0.05 (P=0.0238) | -0.06 (P=0.0122) | -0.07 (P=0.0026) | 0.10  [0.06, 0.11] | <0.0001 | -21 |
| A | -0.07 (P=0.0022) |  |  | 0.06  [0.04, 0.07] | 0.0015 | -10 |
| T |  | -0.09 (P=0.0004) |  | 0.07  [0.03, 0.08] | 0.001 | -10 |
| N |  |  | -0.09 (P=0.0003) | 0.06  [0.03, 0.09] | 0.001 | -10 |

This table shows the results from fitting linear mixed effects models with longitudinal MMSE as outcome and plasma biomarkers added separately or all together to a basic model consisting of age, sex, and education. Beta coefficients are presented in terms of “MMSE points / year per standard deviation change in biomarker value.” R^2^ values were evaluated at the four-year follow-up point, and confidence intervals were calculated using 1000 bootstrapped samples. The basic model consisting of only demographics had R^2^ = 0.04 (95% CI [0.02, 0.05]) and AIC = 4702. Legend: P-values represent an ANOVA comparison to the basic model; AIC_Δ_ values represent the change in AIC compared to the basic model and an AIC_Δ_ value of -2 or lower implies a better fit than the basic model.

**Table S3. Association between CSF biomarkers and longitudinal PACC**

| **Model** | **Beta Coefficient** | | | **R^2^**  **[95% CI]** | **Ref: Basic Model** | |
| --- | --- | --- | --- | --- | --- | --- |
|  | **Plasma**  **Aβ42/Aβ40** | **Plasma**  **P-tau217** | **Plasma**  **NfL** |  | **P value** | **AIC_Δ_** |
| ATN | -0.26 (P<0.0001) | -0.20 (P<0.0001) | -0.20 (P<0.0001) | 0.25  [0.21, 0.28] | <0.0001 | -98 |
| A | -0.25 (P<0.0001) |  |  | 0.13  [0.11, 0.17] | <0.0001 | -30 |
| T |  | -0.28 (P<0.0001) |  | 0.14  [0.12, 0.17] | <0.0001 | -34 |
| N |  |  | -0.30 (P<0.0001) | 0.18  [0.14, 0.21] | <0.0001 | -52 |

This table shows the results from fitting linear mixed effects models with longitudinal PACC as outcome and CSF biomarkers added separately or all together to a basic model consisting of age, sex, and education. Beta coefficients are presented in terms of “PACC points / year per standard deviation change in biomarker value.” R^2^ values were evaluated at the four-year follow-up point, and confidence intervals were calculated using 1000 bootstrapped samples. The basic model consisting of only demographics had R^2^ = 0.07 (95% CI [0.06, 0.11]) and AIC = 6699. Legend: P-values represent an ANOVA comparison to the basic model; AIC_Δ_ values represent the change in AIC compared to the basic model and an AIC_Δ_ value of -2 or lower implies a better fit than the basic model.

**Table S4. Association between plasma biomarkers and conversion to AD dementia with APOE included in basic model**

| **Model** | **Hazard Ratio** | | | **AUC**  **[95% CI]** | **Ref: Basic Model** | |
| --- | --- | --- | --- | --- | --- | --- |
|  | **Plasma**  **Aβ42/Aβ40** | **Plasma**  **P-tau217** | **Plasma**  **NfL** |  | **P value** | **AIC_Δ_** |
| ATN | 1.57 (P=0.0423) | 2.71  (P=0.0021) | 1.09 (P=0.7202) | 0.86  [0.82, 0.93] | 0.0002 | -14 |
| A | 1.69 (P=0.0114) |  |  | 0.81  [0.77, 0.89] | 0.0141 | -4 |
| T |  | 2.99  (P=0.0004) |  | 0.84  [0.75, 0.91] | 0.0001 | -13 |
| N |  |  | 1.47 (P=0.0859) | 0.79  [0.72, 0.89] | 0.0918 | -1 |

This table shows the results from fitting Cox regression models with conversion to AD as outcome and plasma biomarkers added separately or all together to a basic model consisting of age, sex, education, and *APOE* status. Hazard ratios are presented in terms of “increased risk of converting to AD for each standard deviation change in biomarker value.” AUC values were evaluated at the four-year follow-up point, and confidence intervals were calculated using 1000 bootstrapped samples. The basic model consisting of only demographics had AUC = 0.78 (95% CI [0.70, 0.91]) and AIC = 253. Legend: P-values represent an ANOVA comparison to the basic model; AIC_Δ_ values represent the change in AIC compared to the basic model and an AIC_Δ_ value of -2 or lower implies a better fit than the basic model.

**Table S5. Association between plasma biomarkers and conversion to all-cause dementia**

| **Model** | **Hazard Ratio** | | | **AUC**  **[95% CI]** | **Ref: Basic Model** | |
| --- | --- | --- | --- | --- | --- | --- |
|  | **Plasma**  **Aβ42/Aβ40** | **Plasma**  **P-tau217** | **Plasma**  **NfL** |  | **P value** | **AIC_Δ_** |
| ATN | 1.72 (P=0.0019) | 1.93  (P=0.0055) | 1.25 (P=0.2617) | 0.75  [0.68, 0.84] | <0.0001 | -21 |
| A | 1.84 (P=0.0002) |  |  | 0.73  [0.67, 0.82] | 0.0002 | -11 |
| T |  | 2.36  (P=0.0002) |  | 0.72  [0.63, 0.82] | <0.0001 | -14 |
| N |  |  | 1.54 (P=0.0246) | 0.68  [0.60, 0.78] | 0.0274 | -3 |

This table shows the results from fitting Cox regression models with conversion to all-cause dementia as outcome and plasma biomarkers added separately or all together to a basic model consisting of age, sex, and education. Hazard ratios are presented in terms of “increased risk of converting to all-cause dementia for each standard deviation change in biomarker value.” AUC values were evaluated at the four-year follow-up point, and confidence intervals were calculated using 1000 bootstrapped samples. The basic model consisting of only demographics had AUC = 0.66 (95% CI [0.57, 0.75]) and AIC = 368. Legend: P-values represent an ANOVA comparison to the basic model; AIC_Δ_ values represent the change in AIC compared to the basic model and an AIC_Δ_ value of -2 or lower implies a better fit than the basic model.

**Table S6. Power analysis for a theoretical clinical trial**

| **Trial Endpoint** | **Biomarker** | **Sample Size**  **Reduction (%)** | **P-value** |
| --- | --- | --- | --- |
| Change in PACC | Plasma Aβ42/Aβ40 | 45 [20, 63] | 0.0032 |
|  | Plasma P-tau217 | 47 [16, 65] | 0.0072 |
|  | Plasma NfL | 41 [5, 63] | 0.028 |
|  | Combined Model | 70 [54, 81] | < 0.001 |
| Conversion to AD | Plasma Aβ42/Aβ40 | 48 [38, 56] | < 0.001 |
|  | Plasma P-tau217 | 50 [35, 60] | 0.0008 |
|  | Plasma NfL | 24 [-10, 45] | 0.2096 |
|  | Combined Model | 63 [53, 70] | < 0.001 |

This tables shows the reduction in sample size resulting from using plasma biomarkers for inclusion enrichment in theoretical clinical trials aimed at slowing decline in PACC or reducing risk of conversion to AD dementia in a CU population. Sample sizes were estimated for a clinical trial in which pre-defined cutoffs for each biomarker were used as a screening inclusion threshold. Sample size reductions presented in the table are for the enriched trial relative to a trial which does not use any biomarkers for screening/enrichment. Pre-defined threshold values are described in Methods S2. Confidence intervals were derived using 1000 bootstrapped trials.

**Supplementary Figures**

**Figure S1. Relationship between plasma biomarkers**

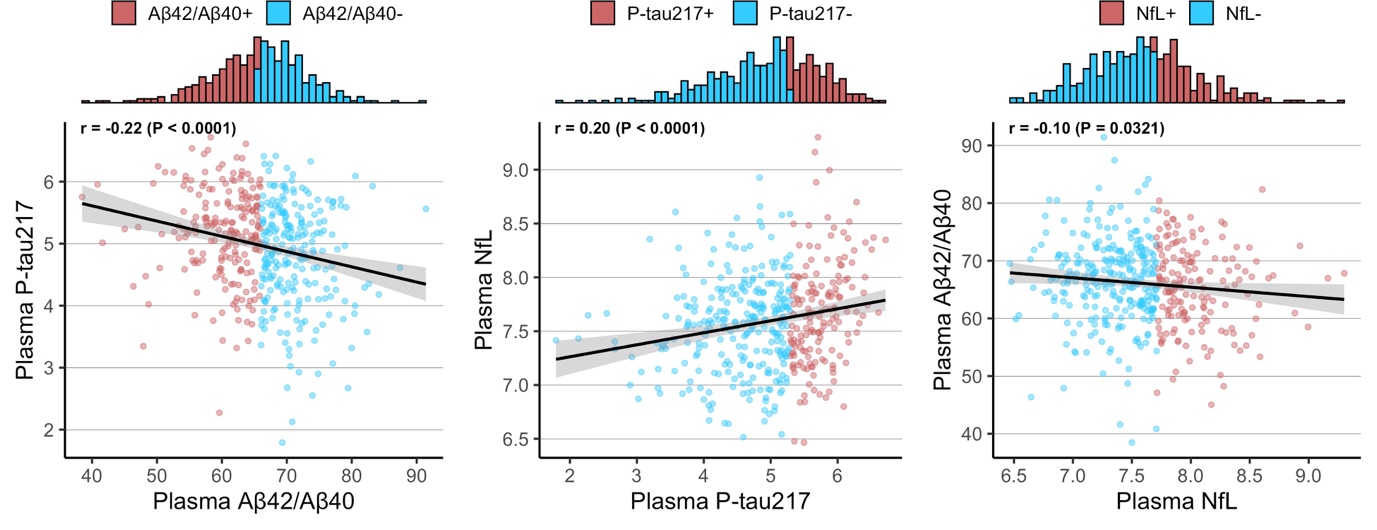

This figure shows the relationship between each pair of plasma biomarkers in the study population. All biomarkers were natural log-transformed and statistical associations were tested using Pearson correlation. The upper panels show the histogram distributions for each corresponding biomarker labelled on the x-axis and each individual is colored based on biomarker status (positive or negative; defined using pre-defined cutoffs) for the biomarker on the x-axis.

**Figure S2. Relationship between plasma biomarkers and change in MMSE**

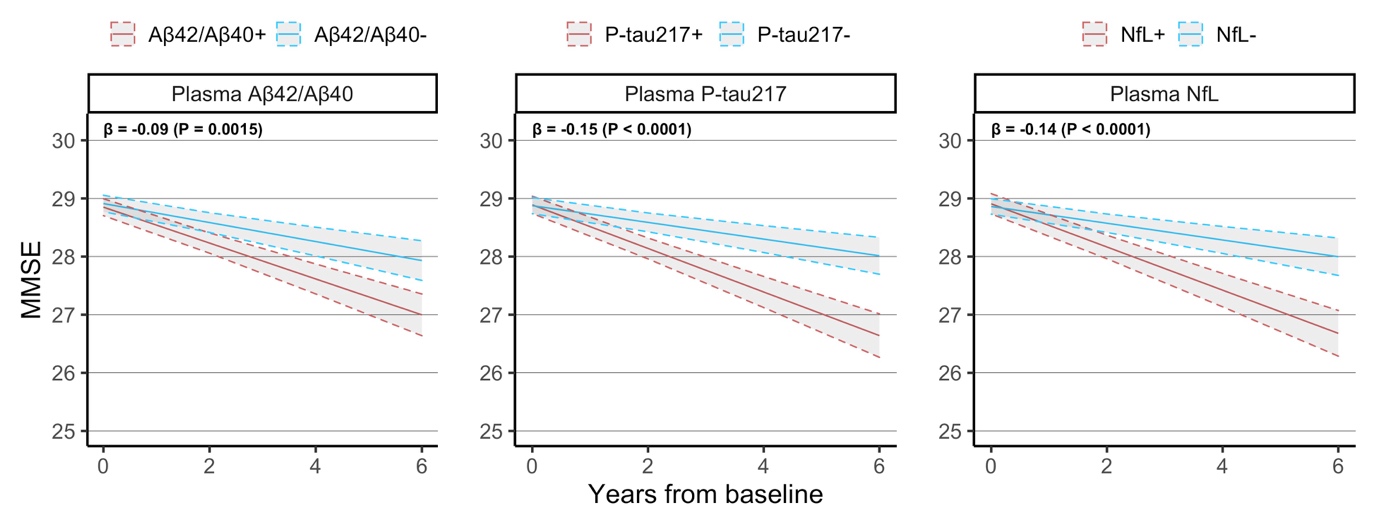

This figure shows the longitudinal MMSE trajectory estimated for a CU individual with average age, average education, female sex and either biomarker-negative or biomarker-positive. Beta coefficients at the top left of each panel are presented in terms of “points / year per standard deviation change in biomarker value” and are derived from linear mixed effects models with longitudinal MMSE as outcome and age, sex, education, plus each plasma biomarker included separately from each other.

**Figure S3. Relationship between plasma biomarkers and conversion to all-cause dementia**

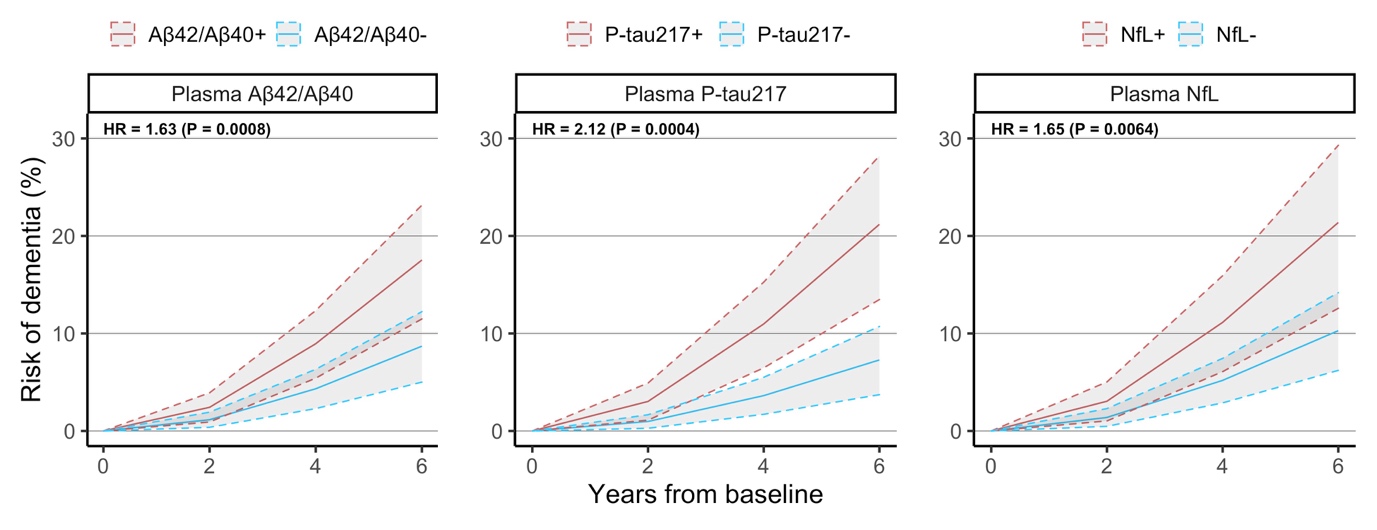

This figure shows the conversion to all-cause dementia estimated for a CU individual with average age, average education, female sex and either biomarker-negative or biomarker-positive. Hazard ratios at the top left of each panel are presented in terms of “increased risk of converting to all-cause dementia per standard deviation change in biomarker value” and are derived from Cox regression models with conversion to all-cause dementia as outcome and age, sex, education, plus each plasma biomarker included separately from each other.
